# Adverse events in both childhood and adulthood are associated with molecular, clinical and functional markers of ageing

**DOI:** 10.1101/2025.06.03.25328874

**Authors:** Monica Aas, Thole H Hoppen, Nexhmedin Morina, Shiyu Zhang, Bin Li, Vid Mlakar, Julian Mutz

## Abstract

Adverse events, including interpersonal trauma, have been linked to poorer health outcomes. However, the biological mechanisms linking adversity and health outcomes remain poorly understood. Here, we investigated associations between molecular, clinical and functional markers of biological ageing and adverse events experienced in childhood and adulthood. Using data from the UK Biobank (*N* = 153,557 middle-aged and older adults), we investigated whether adversity experienced in childhood and/or adulthood was associated with metabolomic ageing, frailty, telomere length and grip strength. Across childhood and adulthood exposures, adversity and its severity were most consistently associated with higher frailty index values, with the strongest associations observed in individuals exposed to multiple types of adverse events. Individuals who experienced adversity in both childhood and adulthood also had a metabolite-predicted age (MileAge) exceeding chronological age and lower grip strength. Abuse was more consistently associated with ageing biomarkers than neglect. Our findings suggest that adversity is associated with older biological ageing profiles across multiple domains, which may be part of the mechanism linking early and later life adversities to poorer health outcomes and premature mortality.

## Introduction

Adverse events occurring in both childhood and adulthood have been linked to detrimental health outcomes in later life (Aas et al., 2017; Alley, Gassen, & Slavich, 2025; Goodwin & Stein, 2004; Sowder, A., & and Fishalow, 2018). Chronic or repeated exposure to adverse events is linked to accelerated biological ageing (Bourassa et al., 2023; Polsky, Rentscher, & Carroll, 2022). However, the mechanisms underlying this relationship remain poorly understood. Recent findings suggest that biological ageing can be accelerated by severe stress exposure but may also be restored upon recovery (Poganik et al., 2023), suggesting that both timing and length of exposure are crucial.

Advances in data analytics and the availability of high-throughput molecular data have enabled the development of diverse markers of biological ageing. A promising approach for developing such markers is metabolomics, which captures the interplay between environmental exposures and genetics. The metabolome bridges the gap between genotype and phenotype (Fiehn, 2002), providing insights into cellular processes linked to health and disease. Indeed, metabolomics can identify actionable targets for intervention (Gieger et al., 2008). However, studies investigating the relationship between exposure to adverse events and metabolomic markers of ageing are lacking.

Frailty is a clinical syndrome characterised by reduced physiological reserve and increased vulnerability to adverse health outcomes, including mortality (Mutz, Choudhury, Zhao, & Dregan, 2022). It is often measured using a frailty index, which quantifies the proportion of accumulated health deficits (e.g., symptoms, signs or diseases) that an individual has. Due to demographic changes towards population ageing, frailty is becoming an emerging global health burden. However, it also represents a potentially modifiable target for intervention to extend healthy life expectancy. A recent study of 862 older adults found greater frailty in those with higher stress levels (Lee et al., 2023), indicating that stress exposure may contribute to frailty.

Telomeres are structures at the ends of chromosomes made of repeating TTAGGG nucleotides (Blackburn, 2001). Their role is to guard DNA from cellular damage. When telomeres become critically short, the risk of apoptosis is increased and proliferation is arrested, thus compromising tissue renewal capacity and function (Blasco, 2007). Telomere length is an indicator of cellular replicative history and shorter telomeres are associated with an older chronological age (Blackburn, 2001). Increasing evidence links childhood adversity to accelerated telomere shortening (Epel et al., 2004; Puterman, Lin, Krauss, Blackburn, & Epel, 2015). However, most studies have been conducted in small samples or clinical populations, limiting generalisability.

Grip strength is well-established functional marker of ageing, predictive of morbidity and mortality (Mutz & Lewis, 2021). A recent study based on 879 older adults reported that stressful life events in childhood were associated with lower muscle strength (Duchowny et al., 2024). The authors proposed that imparied skeletal muscle mitochondrial energetics might represent a potential biological pathway through which early-life adversity may impact health later in life. Nevertheless, prior studies have largely examined single markers of biological ageing or investigated adversity limited to either childhood or adulthood, but rarely both.

Here, we use data from the UK Biobank to investigate associations between exposure to adverse events in childhood and adulthood and molecular, clinical and functional markers of biological ageing. Specifically, we examine associations of adversity with metabolomic age (MileAge) delta, a metabolomic mortality profile, frailty, telomere length and grip strength. Extending prior studies, we evaluate multiple markers of biological ageing in a large well-characterised sample. We further provide new insights into the timing (i.e., childhood, adulthood or both), severity (i.e., overall burden of adversity) and types (e.g., emotional abuse or physical neglect) of adverse events. We hypothesised that individuals exposed to adversity in both childhood and adulthood would have older biological ageing profiles compared with those exposed to adversity only in childhood, only in adulthood, or not at all.

## Methods

### Study population

The UK Biobank recruited over 500,000 individuals from England, Scotland and Wales between 2006 and 2010. At baseline, participants completed health and sociodemographic questionnaires, underwent physical examinations and provided biological samples.

### Adversity and trauma

Exposure to adversity and trauma was assessed via the UK Biobank online mental health questionnaire (MHQ) between 2016 and 2017 (Davis et al., 2020).

Adverse events in childhood were assessed using the Childhood Trauma Screener (Glaesmer et al., 2013), a short version of the Childhood Trauma Questionnaire (Bernstein & Fink, 1998). It comprises five items, with responses ranging from “never true” to “very often true”: “When I was growing up, … 1) I felt loved [emotional neglect]; 2) people in my family hit me so hard that it left me with bruises or marks [physical abuse]; 3) I felt that someone in my family hated me [emotional abuse]; 4) someone molested me sexually [sexual abuse]; and 5) there was someone to take me to the doctor if I needed it [physical neglect].” Individuals were classified as having experienced adversity or trauma in childhood (yes/no) based on the following cut-offs: physical abuse, emotional abuse or sexual abuse (at least “rarely true”); emotional neglect or physical neglect (less than “often”; reverse coded). We also derived a sum score (ranging from 0 to 5), reflecting the overall childhood trauma burden. We further derived a weighted sum score ranging from 0 to 15 (Pitharouli et al., 2021). For this, responses were recoded on a scale from 0 to 3 reflecting the severity of the item (0 = no trauma, 1 = mild trauma, 2 = moderate trauma, 3 = severe trauma), based on severity and frequency of the trauma.

Adverse events in adulthood were assessed using questions adapted from the 2010/2011 British Crime Survey questions to identify victims of crime and domestic violence (Khalifeh, Oram, Trevillion, Johnson, & Howard, 2015). Five questions were asked, with responses ranging from “never true” to “very often true”: “Since I was sixteen, … 1) I have been in a confiding relationship [emotional neglect]; 2) a partner or ex-partner repeatedly belittled me to the extent that I felt worthless [emotional abuse]; 3) a partner or ex-partner deliberately hit me or used violence in any other way [physical abuse]; 4) a partner or ex-partner sexually interfered with me, or forced me to have sex against my wishes [sexual abuse]; 5) there was money to pay the rent or mortgage when I needed it [economic hardship].” Individuals were classified as having experienced adversity or trauma in adulthood (yes/no) based on the following cut-offs: emotional abuse, sexual abuse or physical violence (at least “rarely true”); emotional neglect (“sometimes true” or less; reverse coded); economic hardship (“often” or less; reverse coded) (Davis et al., 2020). We also derived a sum score (ranging from 0 to 5), reflecting overall adulthood trauma burden.

### Metabolomic age (MileAge) delta

Nuclear magnetic resonance (NMR) spectroscopy–derived metabolomic biomarkers were quantified in non-fasting plasma samples using the Nightingale Health platform, which ascertains 249 biomarkers (168 in absolute concentrations and 81 derived ratios) (Würtz et al., 2017). Removal of technical variation was performed using the ‘ukbnmr’ R package (algorithm v2) (Ritchie et al., 2023). In a prior study (Mutz, Iniesta, & Lewis, 2024), we developed a metabolomic ageing clock using a Cubist rule-based regression model. Individual-level age predictions were obtained by aggregating the predictions from the ten test sets of the outer loop of the nested cross-validation to avoid potential overfitting. Metabolomic age (MileAge) delta represents the difference between metabolite-predicted and chronological age, with positive values indicating accelerated biological ageing (Mutz et al., 2024).

### Metabolomic mortality profile score

In a previous study (Zhang et al., 2024), we developed a metabolomic mortality profile score. Complementing the 249 biomarkers provided by the Nightingale Health platform, 76 additional lipid, cholesterol and fatty acid ratios were derived (Ritchie et al., 2023), resulting in a total of 325 biomarkers. A Least Absolute Shrinkage and Selection Operator (LASSO) Cox proportional hazards model predicting all-cause mortality was developed in English and Welsh participants (*N* = 234,553). We derived a metabolomic mortality profile score in Scottish participants (*N* = 15,788) as the linear combination of the 54 biomarkers with non-zero coefficients in the LASSO Cox model weighted by their log hazard ratio for all-cause mortality. Higher scores indicate an elevated mortality risk.

### Frailty index

A frailty index was derived from health deficits reported via touch-screen questionnaires or during nurse-led interviews that met the following criteria: indicators of poor health, more prevalent in older individuals, neither rare nor universal, covering multiple areas of functioning and available for ≥ 80% of participants (Williams, Jylhävä, Pedersen, & Hägg, 2019). The 49 items that met those criteria included cardiometabolic, cranial, immunological, musculoskeletal, respiratory and sensory traits, well-being, infirmity, cancer and pain. Categorical variables were dichotomised (deficit absent = zero; deficit present = one) and ordinal variables were mapped onto a score between zero and one. The sum of deficits was divided by the number of possible deficits, resulting in frailty index scores between zero and one, with higher scores indicating greater levels of frailty (Mutz et al., 2022). Participants with missing data for ≥ 10/49 variables were excluded (Williams et al., 2019).

### Telomere length

Leukocyte telomere length was measured using a quantitative polymerase chain reaction (qPCR) assay that expresses telomere length as the ratio of the telomere repeat copy number (T) relative to a single-copy gene (S) encoding haemoglobin subunit beta (Codd et al., 2022). The T/S ratio is proportional to average telomere length (Lai, Wright, & Shay, 2018). Measurements were adjusted for operational and technical parameters (PCR machine, staff member, enzyme batch, primer batch, temperature, humidity, primer batch × PCR machine, primer batch × staff member, A260/A280 ratio of the DNA sample and A260/A280 ratio squared), log*_e_* transformed and *Z*-standardised.

### Grip strength

Maximal grip strength in whole kilogram force units was measured using a Jamar J00105 hydraulic hand dynamometer (measurement range 0-90 kg) for both hands. We used the maximal grip strength of the participant’s self-reported dominant hand. If no data on handedness were available, we used the highest value.

### Covariates

Potential confounders included chronological age, sex, highest educational or professional qualification, gross annual household income, ethnicity and neighbourhood deprivation assessed via the Townsend deprivation index.

### Exclusion criteria

Individuals with missing data or who responded “prefer not to answer” to any questionnaire items on adverse events were excluded from the derived variables (binary phenotypes and sum scores). For item-specific analyses, only those with missing data or with a “prefer not to answer” response for the specific item were excluded. For categorical covariates, “do not know” responses, “prefer not to answer” responses and missing data were coded as a missing level. For the analyses of MileAge delta, we applied exclusions as per the original study (Mutz et al., 2024): women with possible pregnancy, as metabolite profiles differ during pregnancy; individuals with discordant genetic and self-reported sex; and individuals with missing or outlier metabolite values (4× the interquartile range from the median).

### Statistical analyses

Data processing, analyses and visualisations were performed in R (version 4.3.0). Sample characteristics were summarised using means and standard deviations or counts and percentages. Associations between adverse events (exposures) and MileAge delta, the metabolomic mortality profile score, the frailty index, telomere length and grip strength (outcomes) were estimates using ordinary least squares regression. All outcomes were scaled to have a mean equal to zero and a standard deviation of one, allowing for direct comparison of the association estimates. Telomere length and grip strength were reverse coded prior to analysis. For the analysis of adverse childhood events, we examined a binary (yes/no) exposure definition, unweighted and weighted sum scores as well as an ordinal definition of exposure (based on the unweighted sum score). For adverse events in adulthood, we examined a binary (yes/no) exposure definition, an unweighted sum score and an ordinal definition (based on the sum score). We further examined, for both child and adulthood adversity, whether exposure to multiple traumatic events (two or more) was associated with biological ageing markers. Finally, we examined associations with the cross-classification of child and adulthood adverse events (child and adulthood, childhood only, adulthood only and neither, derived from the above binary definitions). For each outcome, we fitted a minimally adjusted model including chronological age and sex (Model 1) and a fully adjusted model also including education, income, ethnicity and deprivation (Model 2). *P*-values were adjusted for multiple testing using the Benjamini-Hochberg correction method, with a two-tailed test and a false discovery rate of 5%. To investigate whether specific types of adverse events were associated with markers of biological ageing, we further examined associations for each individual item included in the questionnaires. We applied the same modelling strategy reported above.

## Results

### Adverse events in childhood

Amongst 153,557 participants, 63,066 (41%) reported experiencing at least one adverse event during childhood (Table S1). Analytical sample sizes are shown in Table S2; those include individuals with/without adulthood adversity exposure. Childhood adverse events (yes/no) were associated with a metabolite-predicted age exceeding chronological age (*β* = 0.020, 95% CI 0.005-0.035, *p* = 0.018), higher frailty index values (*β* = 0.284, 95% CI 0.276-0.293, *p* < 0.001) and shorter telomeres (*β* = 0.014, 95% CI 0.004-0.024, *p* = 0.015) (Figure 1A).

**Figure 1.**
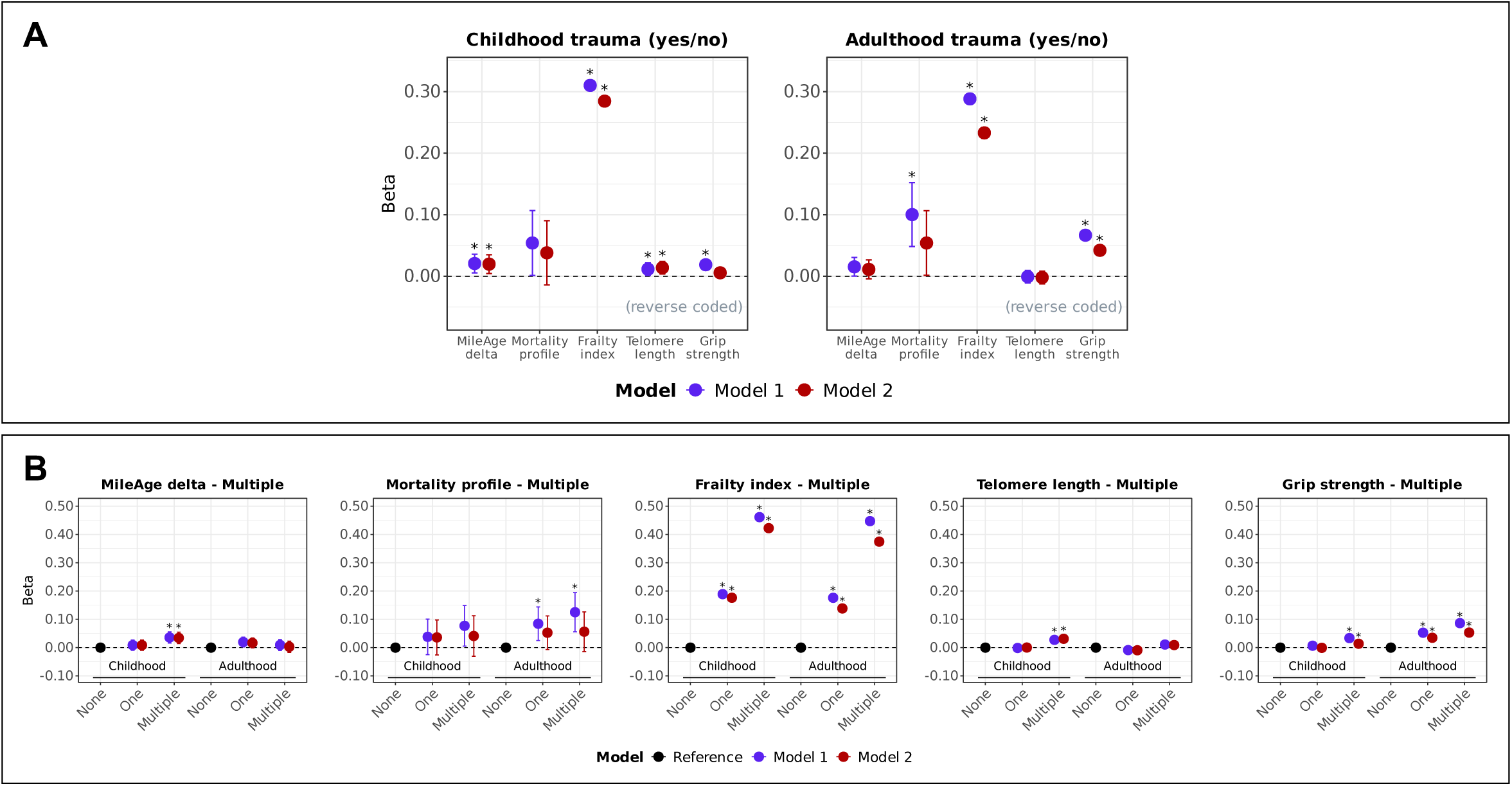
**(A) Ageing markers and trauma (yes/no).** Associations between trauma (adverse/traumatic events in childhood and in adulthood) and ageing markers (MileAge delta, frailty index, telomere length [reverse coded] and grip strength [reverse coded]). Asterisks indicate statistically significant associations, after correcting *p*-values for multiple testing using the Benjamini–Hochberg procedure (across ageing markers and models, separately for childhood and adulthood exposures). **(B) Ageing markers and multiple traumas.** Associations between multiple traumas (adverse/traumatic events in childhood and in adulthood) and ageing markers (MileAge delta, frailty index, telomere length [reverse coded] and grip strength [reverse coded]). Asterisks indicate statistically significant associations, after correcting *p*-values for multiple testing using the Benjamini–Hochberg procedure (across exposure levels and models, separately for each ageing marker and childhood and adulthood exposures). **(A-B)** Estimates shown are ordinary least squares regression beta coefficients and 95% confidence intervals. Model 1– adjusted for chronological age and sex; Model 2–adjusted for chronological age, sex, ethnicity, highest educational/professional qualification, annual gross household income and Townsend deprivation index. Sample sizes reported in Tables S2 and S7.

### Dose-response associations with adversity in childhood

Exposure to multiple (two or more) adverse events in childhood was associated with differences in all biological ageing markers except the metabolomic mortality profile (Figure 1B; Tables S3). Analyses using both unweighted and weighted sum scores indicated that greater adversity burden was associated with older biological ageing profiles across markers, except the metabolomic mortality profile (Table 2). The strongest associations were observed for the frailty index (unweighted: *β* = 0.164, 95% CI 0.160-0.168, *p* < 0.001; weighted: *β* = 0.088, 95% CI 0.086-0.091, *p* < 0.001) (Figure 2A). Although two, three or five adverse events were nominally associated with a MileAge exceeding chronological age (Table S4), only sum scores of five remained statistically significant after multiple testing corrections (*β* = 0.170, 95% CI 0.073-0.267, *p* = 0.003). There was no evidence that any number of adverse events was associated with the metabolomic mortality profile. There was a strong linear association between the number of adverse events and higher frailty index values (*β* = 0.177, 95% CI 0.166-0.187, *p* < 0.001 to *β* = 0.925, 95% CI 0.869-0.981, *p* < 0.001) (Figure 2B). Exposure to two or more or three or more adverse events was associated with shorter telomeres and lower grip strength after full adjustment.

**Figure 2.**
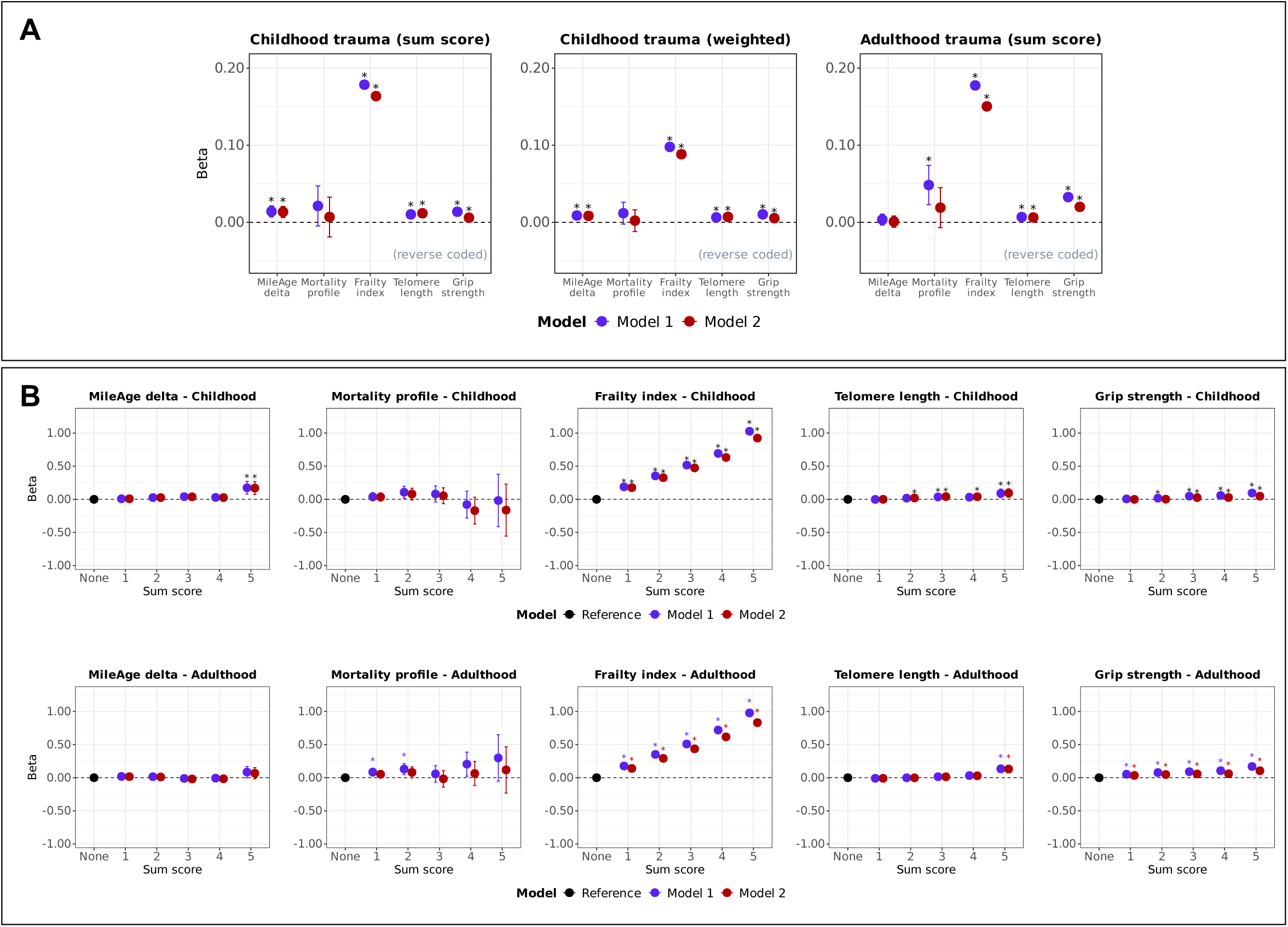
**(A) Ageing markers and trauma (sum scores).** Associations between trauma (adverse/traumatic events in childhood [unweighted and weighted sum scores] and in adulthood) and ageing markers (MileAge delta, frailty index, telomere length [reverse coded] and grip strength [reverse coded]). Asterisks indicate statistically significant associations, after correcting *p*-values for multiple testing using the Benjamini–Hochberg procedure (across ageing markers and models, separately for each sum score). **(B) Ageing markers and trauma (sum scores, categorical).** Associations between trauma (adverse/traumatic events in childhood and in adulthood) and ageing markers (MileAge delta, frailty index, telomere length [reverse coded] and grip strength [reverse coded]). Asterisks indicate statistically significant associations, after correcting *p*-values for multiple testing using the Benjamini–Hochberg procedure (across exposure levels and models, separately for each ageing marker and childhood and adulthood exposures). **(A-B)** Estimates shown are ordinary least squares regression beta coefficients and 95% confidence intervals. Model 1–adjusted for chronological age and sex; Model 2–adjusted for chronological age, sex, ethnicity, highest educational/professional qualification, annual gross household income and Townsend deprivation index. Sample sizes reported in Tables S2 and S7.

### Specific types of adverse events in childhood

Childhood emotional abuse, but not neglect, was associated with higher MileAge delta (*β* = 0.036, 95% CI 0.015-0.056, *p* = 0.002) (Table S5). No statistically significant associations with the metabolomic mortality profile were observed after multiple testing corrections. All types of childhood adversity were associated with higher frailty index scores, and, except for physical neglect, with shorter telomeres (Figure 4). Most childhood adverse events were associated with lower grip strength; however, sexual abuse was not statistically significant after full adjustment, and physical abuse was associated with higher grip strength (*β* = −0.022, 95% CI −0.030 to −0.014, *p* < 0.001).

**Figure 3.**
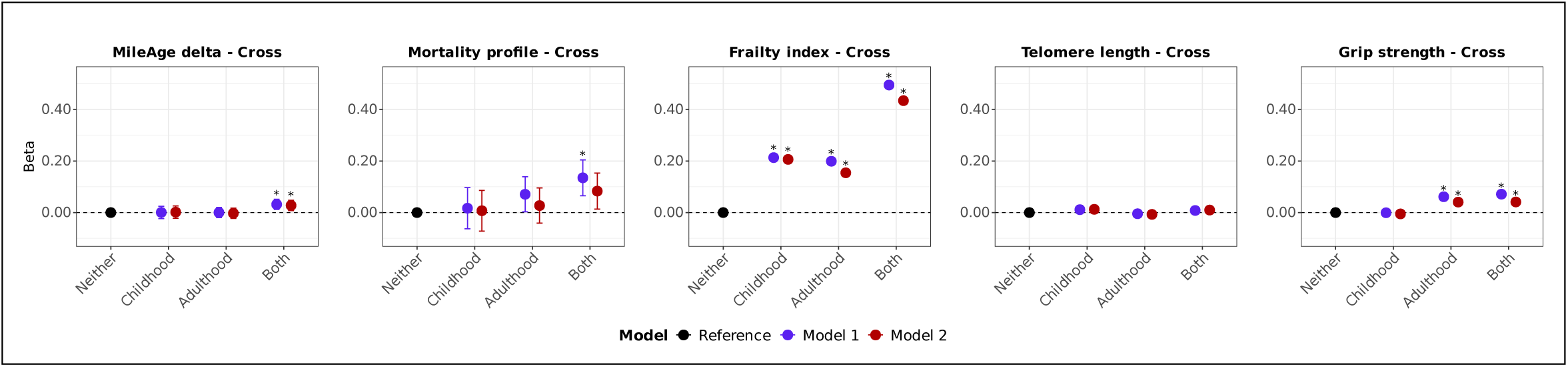
Ageing markers and trauma (cross-classification child and adulthood). Associations between trauma (adverse/traumatic events in childhood and/or adulthood) and ageing markers (MileAge delta, frailty index, telomere length [reverse coded] and grip strength [reverse coded]). Estimates shown are ordinary least squares regression beta coefficients and 95% confidence intervals. Model 1–adjusted for chronological age and sex; Model 2–adjusted for chronological age, sex, ethnicity, highest educational/professional qualification, annual gross household income and Townsend deprivation index. Asterisks indicate statistically significant associations, after correcting *p*-values for multiple testing using the Benjamini–Hochberg procedure (across exposure levels and models, separately for each ageing marker). Sample sizes reported in Table S11.

**Figure 4.**
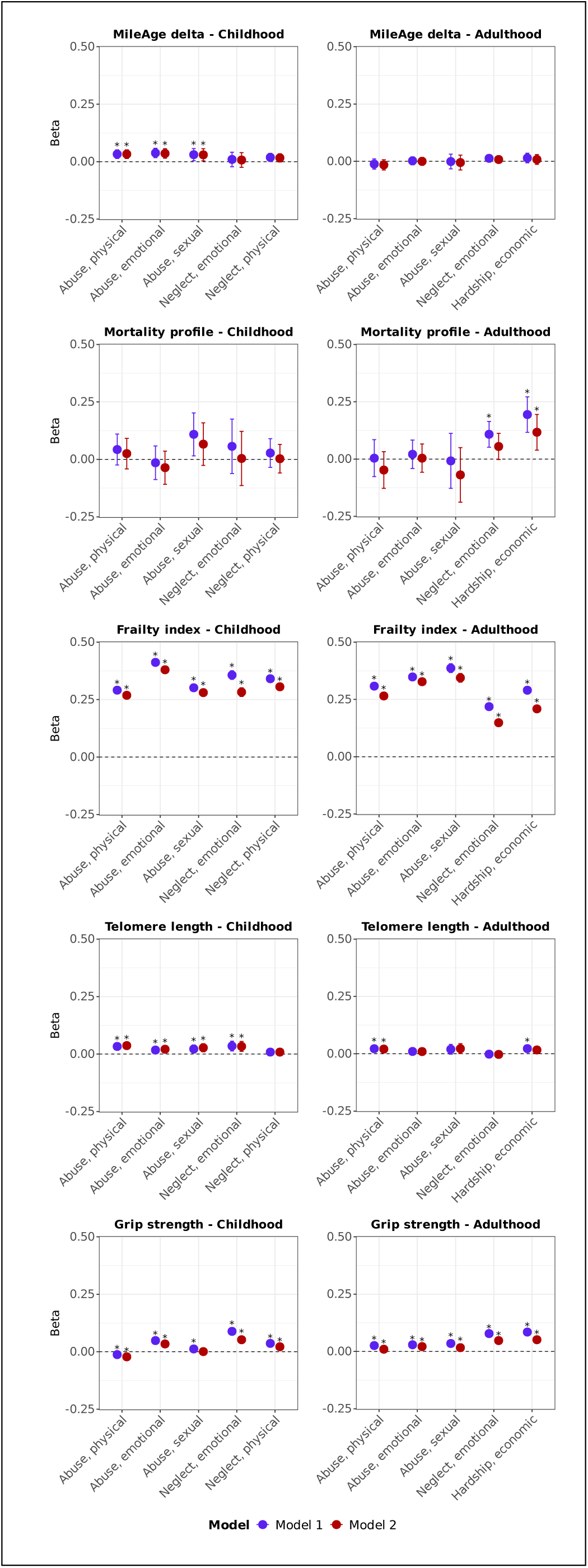
Ageing markers and trauma (item-specific). Associations between specific trauma items (adverse/traumatic events in childhood and in adulthood) and ageing markers (MileAge delta, frailty index, telomere length [reverse coded] and grip strength [reverse coded]). Estimates shown are ordinary least squares regression beta coefficients and 95% confidence intervals. Model 1–adjusted for chronological age and sex; Model 2–adjusted for chronological age, sex, ethnicity, highest educational/professional qualification, annual gross household income and Townsend deprivation index. Asterisks indicate statistically significant associations, after correcting *p*-values for multiple testing using the Benjamini–Hochberg procedure (across trauma items and models, separately for each ageing marker and childhood and adulthood exposures). Sample sizes reported in Tables S2 and S7.

### Adverse events in adulthood

Amongst 150,848 participants, 80,895 (53%) reported at least one adverse event during adulthood (Table S6). Analytical sample sizes are reported in Table S7; those include individuals with/without childhood adversity exposure Adulthood adversity (yes/no) was associated with higher frailty index values (*β* = 0.233, 95% CI 0.224-0.242, *p* < 0.001) and lower grip strength (*β* = 0.042, 95% CI 0.035-0.049, *p* < 0.001) (Figure 1A). The association between adulthood adverse events and higher metabolomic mortality profile scores did not survive multiple testing corrections after full adjustment (*p* = 0.065). No associations were found with MileAge delta or telomere length (Table 1).

**Table 1.**
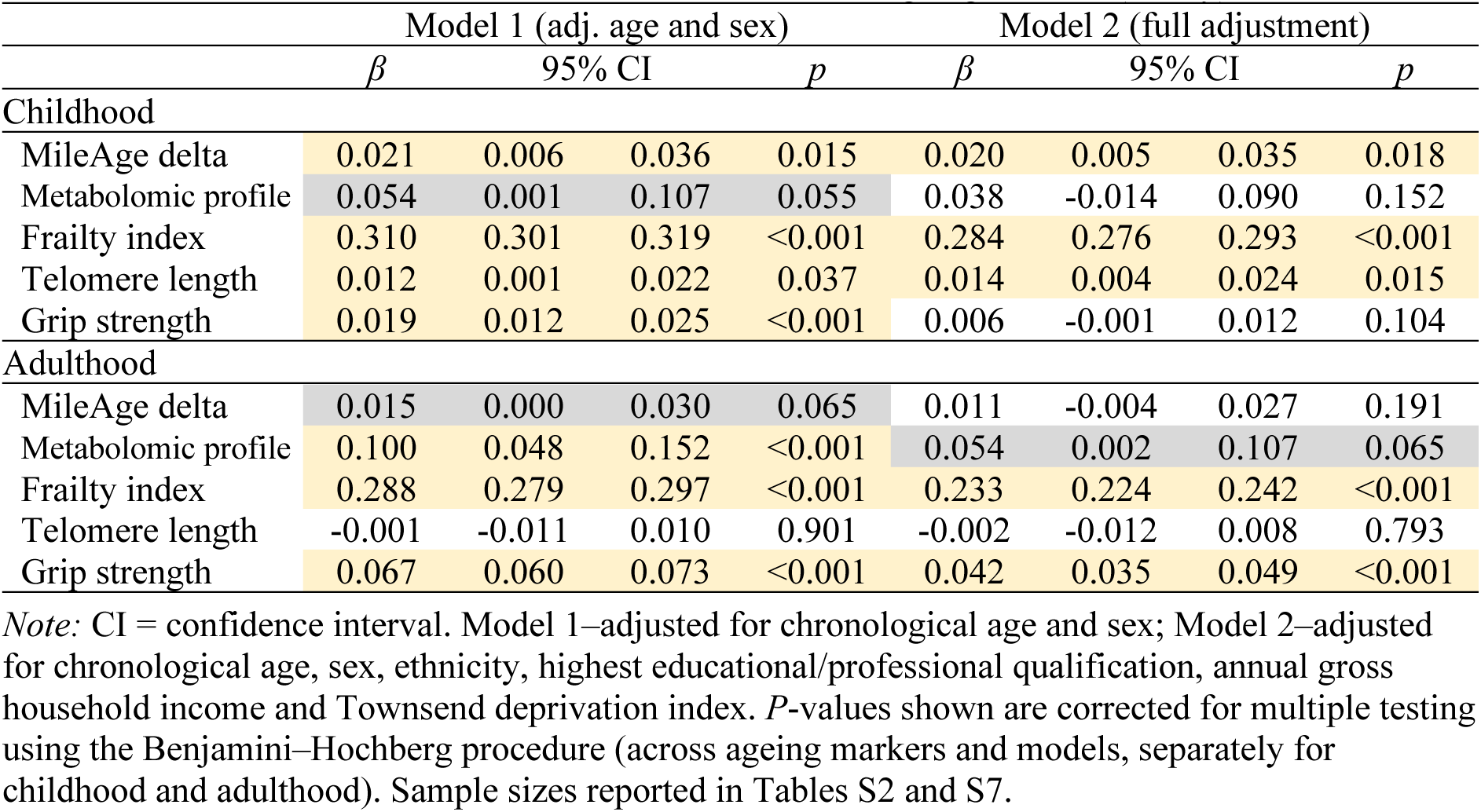
Associations between adverse/traumatic events and ageing markers (binary)

### Dose-response associations with adversity in adulthood

Exposure to multiple adverse events in adulthood was associated with higher metabolomic mortality profile scores, higher frailty index values and lower grip strength (Figure 1B). However, the associations with the metabolomic mortality profile were not statistically significant after full adjustment (Tables S8). Analyses of unweighted sum scores indicated that greater adversity burden in adulthood was associated with all biological ageing markers except MileAge delta (*p* = 0.796) and with the metabolomic mortality profile only in the age and sex-adjusted model (Table 2). The strongest association was observed for the frailty index (*β* = 0.150, 95% 0.146-0.155, *p* < 0.001) (Figure 2A). There was no evidence that any number of adverse events was associated with MileAge delta after multiple testing correction (Table S9). Sum scores of one or two were associated with higher metabolomic mortality profile scores after multiple testing correction in the age and sex-adjusted model (*β* = 0.085, 95% CI 0.025-0.144, *p* = 0.027 and *β* = 0.131, 95% CI 0.048-0.213, *p* = 0.020). A strong linear association between the number of adverse events and higher frailty index values was observed (*β* = 0.140, 95% CI 0.130-0.150, *p* < 0.001 to *β* = 0.831, 95% CI 0.780-0.882, *p* < 0.001) (Figure 2B). A sum score of five was associated with shorter telomeres (*β* = 0.130, 95% CI 0.072-0.189, *p* < 0.001). A greater number of adverse events was consistently associated with lower grip strength (*β* = 0.105, 95% CI 0.067-0.143, *p* < 0.001 for a sum score of five).

**Table 2.** Associations between adverse/traumatic events and ageing markers (sum score) Model 1 (adj. age and sex) Model 2 (full adjustment)

|  | Model 1 (adj. age and sex) |  |  |  | Model 2 (full adjustment) |  |  |  |
| --- | --- | --- | --- | --- | --- | --- | --- | --- |
| | $\beta$ | 95% CI | | $p$ | $\beta$ | 95% CI | | $p$ |
| Childhood |  |  |  |  |  |  |  |  |
| MileAge delta | 0.014 | 0.007 | 0.021 | <0.001 | 0.014 | 0.006 | 0.021 | <0.001 |
| Metabolomic profile | 0.021 | -0.005 | 0.047 | 0.119 | 0.007 | -0.019 | 0.033 | 0.598 |
| Frailty index | 0.179 | 0.174 | 0.183 | <0.001 | 0.164 | 0.160 | 0.168 | <0.001 |
| Telomere length | 0.010 | 0.006 | 0.015 | <0.001 | 0.012 | 0.007 | 0.017 | <0.001 |
| Grip strength | 0.014 | 0.011 | 0.017 | <0.001 | 0.006 | 0.003 | 0.009 | <0.001 |
| Childhood (weighted) |  |  |  |  |  |  |  |  |
| MileAge delta | 0.009 | 0.005 | 0.013 | <0.001 | 0.008 | 0.005 | 0.012 | <0.001 |
| Metabolomic profile | 0.012 | -0.002 | 0.026 | 0.110 | 0.002 | -0.012 | 0.016 | 0.753 |
| Frailty index | 0.098 | 0.096 | 0.100 | <0.001 | 0.088 | 0.086 | 0.091 | <0.001 |
| Telomere length | 0.006 | 0.004 | 0.009 | <0.001 | 0.007 | 0.005 | 0.010 | <0.001 |
| Grip strength | 0.010 | 0.009 | 0.012 | <0.001 | 0.005 | 0.004 | 0.007 | <0.001 |
| Adulthood |  |  |  |  |  |  |  |  |
| MileAge delta | 0.004 | -0.003 | 0.011 | 0.352 | 0.001 | -0.006 | 0.008 | 0.796 |
| Metabolomic profile | 0.048 | 0.023 | 0.074 | <0.001 | 0.019 | -0.007 | 0.045 | 0.187 |
| Frailty index | 0.178 | 0.173 | 0.182 | <0.001 | 0.150 | 0.146 | 0.155 | <0.001 |
| Telomere length | 0.007 | 0.002 | 0.012 | 0.008 | 0.006 | 0.001 | 0.011 | 0.016 |
| Grip strength | 0.033 | 0.030 | 0.036 | <0.001 | 0.020 | 0.017 | 0.023 | <0.001 |

*Note:* CI = confidence interval. Model 1–adjusted for chronological age and sex; Model 2–adjusted for chronological age, sex, ethnicity, highest educational/professional qualification, annual gross household income and Townsend deprivation index. *P*-values shown are corrected for multiple testing using the Benjamini–Hochberg procedure (across ageing markers and models, separately for each sum score). Sample sizes reported in Tables S2 and S7.

### Specific types of adverse events in adulthood

There was no evidence that any type of adulthood adverse event was associated with MileAge delta (Table S10). Emotional neglect and economic hardship were associated with higher metabolomic mortality profile scores, but only economic hardship remained statistically significant after full adjustment (*β* = 0.117, 95% CI 0.039-0.195, *p* = 0.011). All adulthood adverse events were associated with higher frailty index values (Figure 4). These associations were stronger for abuse than for emotional neglect or economic hardship. Only physical abuse remained associated with shorter telomeres after full adjustment and multiple testing correction (*β* = 0.020, 95% CI 0.005-0.035, *p* = 0.025). All adulthood adversity types were associated with lower grip strength.

### Cross-classification of adverse events in childhood and adulthood

A subset of 40,086 (27.1%) individuals experienced adverse events in both childhood and adulthood (Table S11). Exposure to adversity in childhood only, adulthood only, or in both childhood and adulthood was associated with higher frailty index values (Figure 3). Associations were stronger in childhood than adulthood adversity (e.g., *β* = 0.206, 95% CI 0.192-0.220, *p* < 0.001 versus *β* = 0.154, 95% CI 0.143-0.166, *p* < 0.001 for the frailty index, respectively) and greatest amongst individuals exposed to both (*β* = 0.434, 95% CI 0.422-0.445, *p* < 0.001). Only individuals exposed to adversity in both childhood and adulthood had a metabolite-predicted age exceeding chronological age (*β* = 0.028, 95% CI 0.008-0.048, *p* = 0.021) (Table S12). No associations with the metabolomic mortality profile were statistically significant after full adjustment and multiple testing corrections. Exposure to adverse events in adulthood only or in both childhood and adulthood was associated with lower grip strength, of comparable magnitude. No other associations between adversity and ageing markers (telomere length, metabolite-predicted age, metabolomic mortality profile or grip strength) survived corrections for multiple testing. An overview of all results is shown in Table S13.

## Discussion

In this large, well-characterised sample of middle-aged and older adults, we investigated whether adverse or traumatic events experienced in childhood and/or adulthood were associated with molecular, clinical and functional markers of biological ageing. We found that individuals who experienced both childhood and adulthood adversity had a metabolite-predicted age (MileAge) exceeding their chronological age, greater frailty and lower grip strength compared to those who experienced adversity in only one life stage or not at all. Associations with shorter telomeres were observed for childhood adversity, particularly experiences of abuse.

Early life adversities may exert long-lasting effects on health, potentially through mechanisms such as epigenetic modifications, more so than later life adverse events (Meaney, 2010). In relation to adversity and mental health, the timing of childhood adverse events is also important (Aas & Etain, 2020; Andersen & Teicher, 2008). Windows of vulnerability potentially occur during periods of rapid development, and synaptic pruning during childhood and adolescence might unmask underlying predispositions. However, our findings suggest that for biological ageing markers, both early life as well as later life adversity are associated with worse outcomes. This finding aligns with prior research suggesting that cumulative trauma exposure exerts a greater impact on the development of psychopathology (Degenhardt et al., 2022; Ogle, Rubin, & Siegler, 2014). Our study identified mostly older biological ageing profiles in individuals with more adverse events in childhood and in adulthood, apart from physical abuse, which was associated with greater grip strength in adulthood. We speculate that this association reflects a compensatory defensive mechanism in adults experiencing physical abuse.

Previously, we reported shorter telomeres in adults with a history of childhood adversity, primarily in clinical cohorts with severe mental disorders (Aas et al., 2019). In contrast, here we analysed data from a large community-based sample. Compared to the general population, individuals with severe mental disorders are more likely to report adversities (Etain & Aas, 2021; Varese et al., 2012). Both in the current and our prior study, telomere length was measured using qPCR, a validated method for assessing average telomere length (Lindrose et al., 2021). However, qPCR does not capture the number of critically short telomeres or cell-specific variation. Our findings suggest that the frailty index and MileAge delta may better capture the possible impact of adversity on biological ageing.

Our findings hold significant relevance for mental health research and practice, as they reinforce a robust body of evidence linking trauma exposure across the lifespan to poor mental health outcomes. Consistent with previous literature, exposure to adverse events in both childhood and adulthood is associated with increased vulnerability to conditions such as depression and PTSD (Hoppen, Stefan Priebe, Vetter, & Morina, 2021; Kessler et al., 2017). Future research should further investigate how molecular, clinical and functional markers of biological ageing—such as those assessed here—relate to mental health outcomes in individuals exposed to trauma. Such work may help elucidate the interaction between biological and psychological factors and their combined influence on overall health and well-being.

Certain limitations are worth noting. First, the cross-sectional study design precludes causal inference. Future studies could expand on our findings and investigate the links between adversity and changes in metabolomic ageing, frailty, telomere attrition and functional decline prospectively. Second, the current study was performed in a community-based population and not, for instance, exclusively in people with severe mental disorders who have a shorter life expectancy on average (Chang et al., 2023) and are more likely to experience higher levels of adversity (Aas & Etain, 2020; Etain & Aas, 2021; Pruessner, Cullen, Aas, & Walker, 2017). Third, whilst qPCR is often employed in epidemiological research, due to its ability to provide results using only a small amount of DNA (Lindrose et al., 2021), a better estimate of the relationship between telomeres and adversity could have been achieved using a measurement technique with a higher sensitivity to short telomeres. Fourth, the data on adverse events were collected as part of the MHQ follow-up between 2016 and 2017. The possibility of reverse causality between adverse events in adulthood and biological ageing markers can therefore not be excluded. Finally, we used a short version of the CTQ, and thus, may have missed details about childhood adverse events not captured in this version.

## Conclusion

Both childhood and adulthood adverse events were negatively associated with molecular, clinical and functional markers of biological ageing. Accelerated biological ageing across diverse pathways might form part of the mechanism linking early and later life adversities to poorer long-term health outcomes and lifespan.

## Data Availability

The data used are available to all bona fide researchers for health-related research that is in the public interest, subject to an application process and approval criteria. Study materials are publicly available online at http://www.ukbiobank.ac.uk.

## Acknowledgements

MA is funded by the MRC fellowship (#MR/W027720/1). JM is funded by the King’s Prize Fellowship. Computational analyses were supported by King’s Computational Research, Engineering and Technology Environment (CREATE). This research has been conducted using data from UK Biobank. Data access permission has been granted under UK Biobank application 45514 (PI: JM).

## Financial disclosures

The authors declare no conflicts of interest.

## Authorship contributions

MA and JM conceived the idea of the study. JM acquired the data and performed the statistical analysis. SZ and BL developed the metabolomic mortality profile score. MA and JM wrote the manuscript. MA, THH, NM and JM interpreted the findings and revised the manuscript. All authors read and approved the final manuscript.

## Ethics

Ethical approval for the UK Biobank study has been granted by the National Information Governance Board for Health and Social Care and the NHS North West Multicentre Research Ethics Committee (11/NW/0382). No project-specific ethical approval is needed.

## Data sharing statement

The data used are available to all *bona fide* researchers for health-related research that is in the public interest, subject to an application process and approval criteria. Study materials are publicly available online at http://www.ukbiobank.ac.uk.

## Supplementary material

Supplementary information is available online.

## Notes

### Competing Interest Statement

The authors have declared no competing interest.

### Summary of Updates

Fixed citation in-text and in reference list for Hoppen, T. H., Stefan Priebe, Vetter, I., & Morina, N. (2021). Global burden of post-traumatic stress disorder and major depression in countries affected by war between 1989 and 2019: a systematic review and meta-analysis. BMJ Global Health, 6(7), e006303. doi:10.1136/bmjgh-2021-006303

