## Supplementary material for "Adverse events in both childhood and adulthood are associated with molecular, clinical and functional markers of ageing"

This material accompanies the article

### 1. Sample characteristics childhood trauma

**Table S1.** Sample characteristics stratified by childhood trauma

| | Childhood trauma | | |
|---|---|---|---|
| | No (*N*=90,491) | Yes (*N*=63,066) | Full sample (*N*=153,557) |
| MileAge delta, mean (SD)[1] | -0.06 (3.74) | 0.01 (3.80) | -0.03 (3.76) |
| Mortality profile, mean (SD)[1] | -45.11 (0.52) | -45.10 (0.52) | -45.11 (0.52) |
| Frailty index, mean (SD)[1] | 0.10 (0.06) | 0.12 (0.07) | 0.11 (0.07) |
| T/S ratio, mean (SD)[1] | 0.05 (0.99) | 0.06 (0.99) | 0.05 (0.99) |
| Grip strength, mean (SD)[1] | 32.43 (10.95) | 32.48 (11.13) | 32.45 (11.03) |
| Age, mean (SD) | 56.86 (7.67) | 55.76 (7.77) | 56.41 (7.73) |
| Sex | | | |
| Female | 50894 (56.2%) | 35578 (56.4%) | 86472 (56.3%) |
| Male | 39597 (43.8%) | 27488 (43.6%) | 67085 (43.7%) |
| Ethnicity | | | |
| White | 88549 (97.9%) | 60189 (95.4%) | 148738 (96.9%) |
| Mixed | 292 (0.3%) | 500 (0.8%) | 792 (0.5%) |
| Black | 357 (0.4%) | 710 (1.1%) | 1067 (0.7%) |
| Asian | 570 (0.6%) | 691 (1.1%) | 1261 (0.8%) |
| Chinese | 118 (0.1%) | 231 (0.4%) | 349 (0.2%) |
| Other | 344 (0.4%) | 493 (0.8%) | 837 (0.5%) |
| Missing[2] | 261 (0.3%) | 252 (0.4%) | 513 (0.3%) |
| Highest qualification | | | |
| None | 5922 (6.5%) | 4522 (7.2%) | 10444 (6.8%) |
| O levels/GCSEs/CSEs | 21113 (23.3%) | 14782 (23.4%) | 35895 (23.4%) |
| A levels/NVQ/HND/HNC[3] | 21047 (23.3%) | 15030 (23.8%) | 36077 (23.5%) |
| Degree | 41588 (46.0%) | 28120 (44.6%) | 69708 (45.4%) |
| Missing[2] | 821 (0.9%) | 612 (1.0%) | 1433 (0.9%) |
| Household income[4] | | | |
| Very low | 10056 (11.1%) | 8769 (13.9%) | 18825 (12.3%) |
| Low | 18971 (21.0%) | 13290 (21.1%) | 32261 (21.0%) |
| Medium | 23519 (26.0%) | 16597 (26.3%) | 40116 (26.1%) |
| High | 21818 (24.1%) | 14380 (22.8%) | 36198 (23.6%) |
| Very high | 6804 (7.5%) | 4302 (6.8%) | 11106 (7.2%) |
| Missing[2] | 9323 (10.3%) | 5728 (9.1%) | 15051 (9.8%) |
| Townsend deprivation | | | |
| Q1 | 21842 (24.1%) | 12961 (20.6%) | 34803 (22.7%) |
| Q2 | 20451 (22.6%) | 12599 (20.0%) | 33050 (21.5%) |
| Q3 | 18965 (21.0%) | 12673 (20.1%) | 31638 (20.6%) |
| Q4 | 16964 (18.7%) | 13119 (20.8%) | 30083 (19.6%) |
| Q5 | 12162 (13.4%) | 11626 (18.4%) | 23788 (15.5%) |
| Missing[2] | 107 (0.1%) | 88 (0.1%) | 195 (0.1%) |

*Note:* Numbers shown are counts and percentages unless indicated otherwise. SD = standard deviation. GCSEs = general certificate of secondary education; CSE = certificate of secondary education; NVQ = national vocational qualification; HND = higher national diploma; HNC = higher national certificate. [1] Sample sizes for health and ageing markers: $n$ = 69,451 (MileAge delta), 4260 (mortality profile), 153,387 (frailty index), 144,968 (T/S ratio) and 153,055 (grip strength). [2] Missing data may also include “do not know” or “prefer not to answer”. [3] Also includes 'other professional qualifications'. [4] Annual household income groups: very low (<£18,000), low (£18,000–£30,999), middle (£31,000–£51,999), high (£52,000–£100,000) and very high (>£100,000).

#### 2. Analytical sample sizes childhood trauma

**Table S2.** Childhood trauma analytical sample sizes

| Childhood trauma | MileAge delta | Mortality profile | Frailty index | Telomere length | Grip strength |
|---|---|---|---|---|---|
| No | 41138 (59.2%) | 2584 (60.7%) | 90412 (58.9%) | 85479 (59.0%) | 90201 (58.9%) |
| Yes | 28313 (40.8%) | 1676 (39.3%) | 62975 (41.1%) | 59489 (41.0%) | 62854 (41.1%) |
| 0 | 41138 (59.2%) | 2584 (60.7%) | 90412 (58.9%) | 85479 (59.0%) | 90201 (58.9%) |
| 1 | 15800 (22.7%) | 987 (23.2%) | 34869 (22.7%) | 32925 (22.7%) | 34813 (22.7%) |
| 2 | 7068 (10.2%) | 397 (9.3%) | 15853 (10.3%) | 14994 (10.3%) | 15817 (10.3%) |
| 3 | 3627 (5.2%) | 204 (4.8%) | 8161 (5.3%) | 7719 (5.3%) | 8148 (5.3%) |
| 4 | 1404 (2.0%) | 70 (1.6%) | 3181 (2.1%) | 2987 (2.1%) | 3169 (2.1%) |
| 5 | 414 (0.6%) | 18 (0.4%) | 911 (0.6%) | 864 (0.6%) | 907 (0.6%) |
| None | 41138 (59.2%) | 2584 (60.7%) | 90412 (58.9%) | 85479 (59.0%) | 90201 (58.9%) |
| One | 15800 (22.7%) | 987 (23.2%) | 34869 (22.7%) | 32925 (22.7%) | 34813 (22.7%) |
| Multiple | 12513 (18.0%) | 689 (16.2%) | 28106 (18.3%) | 26564 (18.3%) | 28041 (18.3%) |
| Sum score[1] | 0.70 (1.04) | 0.65 (0.99) | 0.71 (1.05) | 0.71 (1.05) | 0.71 (1.05) |
| Weighted score[1] | 1.24 (1.96) | 1.13 (1.83) | 1.25 (1.97) | 1.25 (1.97) | 1.25 (1.97) |
| Specific items | | | | | |
| Abuse, physical | 57400 (81.0%) | 3589 (82.5%) | 127000 (81.0%) | 119995 (81.0%) | 126735 (81.0%) |
| Yes | 13471 (19.0%) | 763 (17.5%) | 29730 (19.0%) | 28102 (19.0%) | 29663 (19.0%) |
| Abuse, emotional | 59969 (84.7%) | 3734 (85.8%) | 132162 (84.4%) | 124906 (84.4%) | 131875 (84.4%) |
| Yes | 10873 (15.3%) | 619 (14.2%) | 24475 (15.6%) | 23102 (15.6%) | 24428 (15.6%) |
| Abuse, sexual | 64195 (91.5%) | 3952 (91.8%) | 141628 (91.2%) | 133820 (91.2%) | 141329 (91.2%) |
| Yes | 5998 (8.5%) | 353 (8.2%) | 13613 (8.8%) | 12874 (8.8%) | 13580 (8.8%) |
| Neglect, emotional | 66581 (94.3%) | 4128 (95.2%) | 147045 (94.2%) | 138942 (94.2%) | 146731 (94.2%) |
| Yes | 4019 (5.7%) | 210 (4.8%) | 8971 (5.8%) | 8487 (5.8%) | 8952 (5.8%) |
| Neglect, physical | 55073 (77.8%) | 3439 (79.1%) | 121258 (77.5%) | 114623 (77.5%) | 120997 (77.5%) |
| Yes | 15689 (22.2%) | 906 (20.9%) | 35203 (22.5%) | 33236 (22.5%) | 35132 (22.5%) |

*Note:* Numbers shown are counts and percentages unless indicated otherwise. [1] mean and standard deviation.

#### 3. Multiple adverse/traumatic events in childhood

**Table S3.** Associations between exposure to multiple adverse/traumatic events in childhood and ageing markers

| | Model 1 (adj. age and sex) | | | | Model 2 (full adjustment) | | | |
|---|---|---|---|---|---|---|---|---|
| | *β* | 95% CI | | *p* | *β* | 95% CI | | *p* |
| MileAge delta | | | | | | | | |
| None | | | | Reference | | | | |
| One | 0.009 | -0.010 | 0.027 | 0.359 | 0.009 | -0.010 | 0.027 | 0.359 |
| Multiple | 0.036 | 0.016 | 0.056 | 0.002 | 0.034 | 0.014 | 0.054 | 0.002 |
| Metabolomic profile | | | | | | | | |
| None | | | | Reference | | | | |
| One | 0.038 | -0.025 | 0.101 | 0.260 | 0.036 | -0.026 | 0.098 | 0.260 |
| Multiple | 0.077 | 0.005 | 0.149 | 0.144 | 0.041 | -0.030 | 0.113 | 0.260 |
| Frailty index | | | | | | | | |
| None | | | | Reference | | | | |
| One | 0.189 | 0.178 | 0.200 | <0.001 | 0.176 | 0.166 | 0.187 | <0.001 |
| Multiple | 0.461 | 0.450 | 0.473 | <0.001 | 0.422 | 0.411 | 0.434 | <0.001 |
| Telomere length | | | | | | | | |
| None | | | | Reference | | | | |
| One | -0.001 | -0.014 | 0.011 | 0.963 | 0.000 | -0.012 | 0.013 | 0.963 |
| Multiple | 0.027 | 0.014 | 0.041 | <0.001 | 0.031 | 0.018 | 0.045 | <0.001 |
| Grip strength | | | | | | | | |
| None | | | | Reference | | | | |
| One | 0.007 | -0.001 | 0.015 | 0.131 | -0.001 | -0.009 | 0.007 | 0.867 |
| Multiple | 0.034 | 0.025 | 0.042 | <0.001 | 0.014 | 0.005 | 0.022 | 0.004 |

*Note:* CI = confidence interval. Model 1–adjusted for chronological age and sex; Model 2–adjusted for chronological age, sex, ethnicity, highest educational/professional qualification, annual gross household income and Townsend deprivation index. *P*-values shown are corrected for multiple testing using the Benjamini–Hochberg procedure (across exposure levels and models, separately for each ageing marker and childhood and adulthood exposures). Sample sizes reported in Table S2.

### 4. Adverse/traumatic events in childhood (ordinal)

**Table S4.** Associations between adverse/traumatic events in childhood and ageing markers (ordinal)

| | Model 1 (adj. age and sex) | | | | Model 2 (full adjustment) | | | |
|---|---|---|---|---|---|---|---|---|
| | *β* | 95% CI | | *p* | *β* | 95% CI | | *p* |
| MileAge delta | | | | | | | | |
| None | | | | Reference | | | | |
| 1 | 0.009 | -0.010 | 0.027 | 0.356 | 0.009 | -0.010 | 0.027 | 0.356 |
| 2 | 0.027 | 0.002 | 0.052 | 0.068 | 0.026 | 0.001 | 0.052 | 0.068 |
| 3 | 0.040 | 0.006 | 0.074 | 0.065 | 0.038 | 0.004 | 0.072 | 0.068 |
| 4 | 0.029 | -0.024 | 0.082 | 0.356 | 0.027 | -0.027 | 0.080 | 0.356 |
| 5 | 0.175 | 0.079 | 0.272 | 0.003 | 0.170 | 0.073 | 0.267 | 0.003 |
| Metabolomic profile | | | | | | | | |
| None | | | | Reference | | | | |
| 1 | 0.038 | -0.025 | 0.101 | 0.422 | 0.036 | -0.026 | 0.098 | 0.422 |
| 2 | 0.107 | 0.016 | 0.197 | 0.207 | 0.080 | -0.010 | 0.169 | 0.324 |
| 3 | 0.081 | -0.041 | 0.203 | 0.422 | 0.054 | -0.066 | 0.175 | 0.493 |
| 4 | -0.079 | -0.282 | 0.124 | 0.493 | -0.170 | -0.371 | 0.031 | 0.324 |
| 5 | -0.018 | -0.414 | 0.378 | 0.931 | -0.162 | -0.554 | 0.229 | 0.493 |
| Frailty index | | | | | | | | |
| None | | | | Reference | | | | |
| 1 | 0.189 | 0.178 | 0.200 | <0.001 | 0.177 | 0.166 | 0.187 | <0.001 |
| 2 | 0.354 | 0.340 | 0.369 | <0.001 | 0.327 | 0.313 | 0.342 | <0.001 |
| 3 | 0.517 | 0.498 | 0.537 | <0.001 | 0.475 | 0.455 | 0.494 | <0.001 |
| 4 | 0.693 | 0.662 | 0.724 | <0.001 | 0.632 | 0.602 | 0.663 | <0.001 |
| 5 | 1.028 | 0.971 | 1.084 | <0.001 | 0.925 | 0.869 | 0.981 | <0.001 |
| Telomere length | | | | | | | | |
| None | | | | Reference | | | | |
| 1 | -0.001 | -0.014 | 0.011 | 0.945 | 0.000 | -0.012 | 0.013 | 0.958 |
| 2 | 0.018 | 0.001 | 0.035 | 0.051 | 0.021 | 0.004 | 0.038 | 0.031 |
| 3 | 0.037 | 0.014 | 0.059 | 0.007 | 0.041 | 0.019 | 0.064 | 0.003 |
| 4 | 0.033 | -0.003 | 0.068 | 0.089 | 0.040 | 0.004 | 0.075 | 0.046 |
| 5 | 0.090 | 0.025 | 0.155 | 0.017 | 0.096 | 0.031 | 0.161 | 0.013 |
| Grip strength | | | | | | | | |
| None | | | | Reference | | | | |
| 1 | 0.007 | -0.001 | 0.015 | 0.123 | -0.001 | -0.009 | 0.007 | 0.873 |
| 2 | 0.018 | 0.007 | 0.029 | 0.003 | 0.003 | -0.008 | 0.013 | 0.715 |
| 3 | 0.048 | 0.034 | 0.063 | <0.001 | 0.027 | 0.012 | 0.041 | <0.001 |
| 4 | 0.059 | 0.036 | 0.082 | <0.001 | 0.027 | 0.005 | 0.050 | 0.031 |
| 5 | 0.095 | 0.053 | 0.138 | <0.001 | 0.047 | 0.005 | 0.090 | 0.040 |

*Note:* CI = confidence interval. Model 1–adjusted for chronological age and sex; Model 2–adjusted for chronological age, sex, ethnicity, highest educational/professional qualification, annual gross household income and Townsend deprivation index. *P*-values shown are corrected for multiple testing using the Benjamini–Hochberg procedure (across exposure levels and models, separately for each ageing marker and childhood and adulthood exposures). Sample sizes reported in Table S2.

#### 5. Item-specific adverse/traumatic events in childhood

**Table S5.** Associations between childhood trauma items and ageing markers

| | Model 1 (adj. age and sex) | | | | Model 2 (full adjustment) | | | |
|---|---|---|---|---|---|---|---|---|
| | *β* | 95% CI | | *p* | *β* | 95% CI | | *p* |
| MileAge delta | | | | | | | | |
| Abuse, physical | 0.033 | 0.014 | 0.052 | 0.002 | 0.033 | 0.014 | 0.052 | 0.002 |
| Abuse, emotional | 0.038 | 0.017 | 0.058 | 0.002 | 0.036 | 0.015 | 0.056 | 0.002 |
| Abuse, sexual | 0.030 | 0.004 | 0.057 | 0.047 | 0.030 | 0.003 | 0.056 | 0.047 |
| Neglect, emotional | 0.010 | -0.022 | 0.041 | 0.609 | 0.007 | -0.025 | 0.039 | 0.661 |
| Neglect, physical | 0.019 | 0.001 | 0.036 | 0.055 | 0.016 | -0.001 | 0.034 | 0.086 |
| Metabolomic profile | | | | | | | | |
| Abuse, physical | 0.043 | -0.024 | 0.110 | 0.642 | 0.025 | -0.041 | 0.091 | 0.655 |
| Abuse, emotional | -0.015 | -0.088 | 0.058 | 0.865 | -0.036 | -0.108 | 0.036 | 0.642 |
| Abuse, sexual | 0.109 | 0.015 | 0.202 | 0.224 | 0.066 | -0.026 | 0.159 | 0.642 |
| Neglect, emotional | 0.057 | -0.062 | 0.175 | 0.642 | 0.004 | -0.114 | 0.122 | 0.947 |
| Neglect, physical | 0.028 | -0.035 | 0.090 | 0.642 | 0.003 | -0.059 | 0.065 | 0.947 |
| Frailty index | | | | | | | | |
| Abuse, physical | 0.291 | 0.280 | 0.302 | <0.001 | 0.269 | 0.258 | 0.280 | <0.001 |
| Abuse, emotional | 0.412 | 0.400 | 0.424 | <0.001 | 0.380 | 0.368 | 0.392 | <0.001 |
| Abuse, sexual | 0.301 | 0.286 | 0.317 | <0.001 | 0.280 | 0.265 | 0.296 | <0.001 |
| Neglect, emotional | 0.357 | 0.338 | 0.376 | <0.001 | 0.283 | 0.264 | 0.301 | <0.001 |
| Neglect, physical | 0.341 | 0.330 | 0.351 | <0.001 | 0.306 | 0.295 | 0.316 | <0.001 |
| Telomere length | | | | | | | | |
| Abuse, physical | 0.033 | 0.020 | 0.046 | <0.001 | 0.037 | 0.024 | 0.050 | <0.001 |
| Abuse, emotional | 0.017 | 0.004 | 0.031 | 0.018 | 0.021 | 0.007 | 0.035 | 0.005 |
| Abuse, sexual | 0.022 | 0.004 | 0.040 | 0.018 | 0.027 | 0.010 | 0.045 | 0.005 |
| Neglect, emotional | 0.035 | 0.014 | 0.056 | 0.004 | 0.033 | 0.012 | 0.055 | 0.005 |
| Neglect, physical | 0.009 | -0.003 | 0.020 | 0.159 | 0.009 | -0.003 | 0.020 | 0.159 |
| Grip strength | | | | | | | | |
| Abuse, physical | -0.012 | -0.021 | -0.004 | 0.004 | -0.022 | -0.030 | -0.014 | <0.001 |
| Abuse, emotional | 0.049 | 0.040 | 0.058 | <0.001 | 0.034 | 0.025 | 0.043 | <0.001 |
| Abuse, sexual | 0.012 | 0.001 | 0.024 | 0.040 | 0.001 | -0.011 | 0.012 | 0.893 |
| Neglect, emotional | 0.089 | 0.075 | 0.103 | <0.001 | 0.052 | 0.038 | 0.066 | <0.001 |
| Neglect, physical | 0.036 | 0.029 | 0.044 | <0.001 | 0.022 | 0.014 | 0.029 | <0.001 |

*Note:* CI = confidence interval. Model 1–adjusted for chronological age and sex; Model 2–adjusted for chronological age, sex, ethnicity, highest educational/professional qualification, annual gross household income and Townsend deprivation index. *P*-values shown are corrected for multiple testing using the Benjamini–Hochberg procedure (across trauma items and models, separately for each ageing marker and childhood and adulthood exposures). Sample sizes reported in Table S2.

### 6. Sample characteristics adulthood trauma

**Table S6.** Sample characteristics stratified by adulthood trauma

| | Adulthood trauma | | |
|---|---|---|---|
| | No (*N*=69,953) | Yes (*N*=80,895) | Full sample (*N*=150,848) |
| MileAge delta, mean (SD)[1] | -0.08 (3.74) | 0.01 (3.78) | -0.03 (3.76) |
| Mortality profile, mean (SD)[1] | -45.12 (0.51) | -45.09 (0.54) | -45.10 (0.52) |
| Frailty index, mean (SD)[1] | 0.10 (0.06) | 0.12 (0.07) | 0.11 (0.07) |
| T/S ratio, mean (SD)[1] | 0.03 (0.99) | 0.07 (0.99) | 0.05 (0.99) |
| Grip strength, mean (SD)[1] | 33.62 (11.13) | 31.51 (10.85) | 32.49 (11.03) |
| Age, mean (SD) | 56.79 (7.59) | 56.01 (7.80) | 56.38 (7.72) |
| Sex | | | |
| Female | 35764 (51.1%) | 49065 (60.7%) | 84829 (56.2%) |
| Male | 34189 (48.9%) | 31830 (39.3%) | 66019 (43.8%) |
| Ethnicity | | | |
| White | 68646 (98.1%) | 77495 (95.8%) | 146141 (96.9%) |
| Mixed | 269 (0.4%) | 520 (0.6%) | 789 (0.5%) |
| Black | 196 (0.3%) | 857 (1.1%) | 1053 (0.7%) |
| Asian | 340 (0.5%) | 876 (1.1%) | 1216 (0.8%) |
| Chinese | 86 (0.1%) | 254 (0.3%) | 340 (0.2%) |
| Other | 221 (0.3%) | 589 (0.7%) | 810 (0.5%) |
| Missing[2] | 195 (0.3%) | 304 (0.4%) | 499 (0.3%) |
| Highest qualification | | | |
| None | 3930 (5.6%) | 6181 (7.6%) | 10111 (6.7%) |
| O levels/GCSEs/CSEs | 15358 (22.0%) | 19816 (24.5%) | 35174 (23.3%) |
| A levels/NVQ/HND/HNC[3] | 16232 (23.2%) | 19169 (23.7%) | 35401 (23.5%) |
| Degree | 33871 (48.4%) | 34902 (43.1%) | 68773 (45.6%) |
| Missing[2] | 562 (0.8%) | 827 (1.0%) | 1389 (0.9%) |
| Household income[4] | | | |
| Very low | 5725 (8.2%) | 12637 (15.6%) | 18362 (12.2%) |
| Low | 13514 (19.3%) | 18106 (22.4%) | 31620 (21.0%) |
| Medium | 18608 (26.6%) | 20953 (25.9%) | 39561 (26.2%) |
| High | 19239 (27.5%) | 16582 (20.5%) | 35821 (23.7%) |
| Very high | 6398 (9.1%) | 4603 (5.7%) | 11001 (7.3%) |
| Missing[2] | 6469 (9.2%) | 8014 (9.9%) | 14483 (9.6%) |
| Townsend deprivation | | | |
| Q1 | 17953 (25.7%) | 16185 (20.0%) | 34138 (22.6%) |
| Q2 | 16470 (23.5%) | 15978 (19.8%) | 32448 (21.5%) |
| Q3 | 14719 (21.0%) | 16357 (20.2%) | 31076 (20.6%) |
| Q4 | 12649 (18.1%) | 16923 (20.9%) | 29572 (19.6%) |
| Q5 | 8085 (11.6%) | 15337 (19.0%) | 23422 (15.5%) |
| Missing[2] | 77 (0.1%) | 115 (0.1%) | 192 (0.1%) |

*Note:* Numbers shown are counts and percentages unless indicated otherwise. SD = standard deviation. GCSEs = general certificate of secondary education; CSE = certificate of secondary education; NVQ = national vocational qualification; HND = higher national diploma; HNC = higher national certificate. [1] Missing data for health and ageing markers: *n* = 68,126 (MileAge delta), 4205 (mortality profile), 150,690 (frailty index), 142,381 (T/S ratio) and 150,362 (grip strength). [2] Missing data may also include “do not know” or “prefer not to answer”. [3] Also includes 'other professional qualifications'. [4] Annual household income groups: very low (<£18,000), low (£18,000–£30,999), middle (£31,000–£51,999), high (£52,000–£100,000) and very high (>£100,000).

### 7. Analytical sample sizes adulthood trauma

**Table S7.** Adulthood trauma analytical sample sizes

| Adulthood trauma | MileAge delta | Mortality profile | Frailty index | Telomere length | Grip strength |
|---|---|---|---|---|---|
| No | 31760 (46.6%) | 2122 (50.5%) | 69905 (46.4%) | 66063 (46.4%) | 69730 (46.4%) |
| Yes | 36366 (53.4%) | 2083 (49.5%) | 80785 (53.6%) | 76318 (53.6%) | 80632 (53.6%) |
| 0 | 31760 (46.6%) | 2122 (50.5%) | 69905 (46.4%) | 66063 (46.4%) | 69730 (46.4%) |
| 1 | 20978 (30.8%) | 1262 (30.0%) | 46673 (31.0%) | 44108 (31.0%) | 46577 (31.0%) |
| 2 | 9476 (13.9%) | 508 (12.1%) | 20979 (13.9%) | 19774 (13.9%) | 20929 (13.9%) |
| 3 | 3747 (5.5%) | 201 (4.8%) | 8387 (5.6%) | 7967 (5.6%) | 8387 (5.6%) |
| 4 | 1635 (2.4%) | 89 (2.1%) | 3604 (2.4%) | 3387 (2.4%) | 3596 (2.4%) |
| 5 | 530 (0.8%) | 23 (0.5%) | 1142 (0.8%) | 1082 (0.8%) | 1143 (0.8%) |
| None | 31760 (46.6%) | 2122 (50.5%) | 69905 (46.4%) | 66063 (46.4%) | 69730 (46.4%) |
| One | 20978 (30.8%) | 1262 (30.0%) | 46673 (31.0%) | 44108 (31.0%) | 46577 (31.0%) |
| Multiple | 15388 (22.6%) | 821 (19.5%) | 34112 (22.6%) | 32210 (22.6%) | 34055 (22.6%) |
| Sum score[1] | 0.89 (1.07) | 0.80 (1.03) | 0.89 (1.07) | 0.89 (1.07) | 0.89 (1.07) |
| Specific items | | | | | |
| Abuse, physical | 61742 (87.2%) | 3859 (88.7%) | 136568 (87.2%) | 129018 (87.2%) | 136267 (87.2%) |
| Yes | 9053 (12.8%) | 492 (11.3%) | 20031 (12.8%) | 18960 (12.8%) | 19996 (12.8%) |
| Abuse, emotional | 53923 (76.2%) | 3399 (78.1%) | 119066 (76.0%) | 112476 (76.0%) | 118795 (76.0%) |
| Yes | 16880 (23.8%) | 953 (21.9%) | 37541 (24.0%) | 35498 (24.0%) | 37474 (24.0%) |
| Abuse, sexual | 66740 (94.3%) | 4141 (95.2%) | 147535 (94.2%) | 139429 (94.2%) | 147215 (94.2%) |
| Yes | 4057 (5.7%) | 210 (4.8%) | 9039 (5.8%) | 8520 (5.8%) | 9027 (5.8%) |
| Neglect, emotional | 47003 (68.0%) | 2994 (70.3%) | 103941 (67.9%) | 98261 (68.0%) | 103692 (67.9%) |
| Yes | 22155 (32.0%) | 1263 (29.7%) | 49038 (32.1%) | 46283 (32.0%) | 48953 (32.1%) |
| Hardship, economic | 59735 (85.4%) | 3778 (87.7%) | 131984 (85.3%) | 124706 (85.3%) | 131687 (85.3%) |
| Yes | 10199 (14.6%) | 528 (12.3%) | 22670 (14.7%) | 21416 (14.7%) | 22634 (14.7%) |

*Note:* Numbers shown are counts and percentages unless indicated otherwise. [1] mean and standard deviation.

#### 8. Multiple adverse/traumatic events in adulthood

**Table S8.** Associations between exposure to multiple adverse/traumatic events in adulthood and ageing markers

| | Model 1 (adj. age and sex) | | | | Model 2 (full adjustment) | | | |
|---|---|---|---|---|---|---|---|---|
| | *β* | 95% CI | | *p* | *β* | 95% CI | | *p* |
| MileAge delta | | | | | | | | |
| None | Reference | | | | | | | |
| One | 0.020 | 0.002 | 0.037 | 0.107 | 0.017 | -0.001 | 0.034 | 0.126 |
| Multiple | 0.009 | -0.010 | 0.029 | 0.461 | 0.003 | -0.017 | 0.023 | 0.760 |
| Metabolomic profile | | | | | | | | |
| None | Reference | | | | | | | |
| One | 0.084 | 0.025 | 0.144 | 0.011 | 0.053 | -0.007 | 0.112 | 0.110 |
| Multiple | 0.125 | 0.056 | 0.195 | 0.002 | 0.056 | -0.014 | 0.127 | 0.116 |
| Frailty index | | | | | | | | |
| None | Reference | | | | | | | |
| One | 0.176 | 0.166 | 0.186 | <0.001 | 0.139 | 0.128 | 0.149 | <0.001 |
| Multiple | 0.447 | 0.436 | 0.459 | <0.001 | 0.375 | 0.363 | 0.386 | <0.001 |
| Telomere length | | | | | | | | |
| None | Reference | | | | | | | |
| One | -0.009 | -0.020 | 0.003 | 0.182 | -0.009 | -0.021 | 0.002 | 0.182 |
| Multiple | 0.011 | -0.002 | 0.024 | 0.182 | 0.009 | -0.004 | 0.022 | 0.182 |
| Grip strength | | | | | | | | |
| None | Reference | | | | | | | |
| One | 0.053 | 0.045 | 0.060 | <0.001 | 0.035 | 0.027 | 0.042 | <0.001 |
| Multiple | 0.087 | 0.078 | 0.095 | <0.001 | 0.053 | 0.045 | 0.062 | <0.001 |

*Note:* CI = confidence interval. Model 1–adjusted for chronological age and sex; Model 2–adjusted for chronological age, sex, ethnicity, highest educational/professional qualification, annual gross household income and Townsend deprivation index. *P*-values shown are corrected for multiple testing using the Benjamini–Hochberg procedure (across exposure levels and models, separately for each ageing marker and childhood and adulthood exposures). Sample sizes reported in Table S7.

### 9. Adverse/traumatic events in adulthood (ordinal)

**Table S9.** Associations between adverse/traumatic events in adulthood and ageing markers (ordinal)

| | Model 1 (adj. age and sex) | | | | Model 2 (full adjustment) | | | |
|---|---|---|---|---|---|---|---|---|
| | *β* | 95% CI | | *p* | *β* | 95% CI | | *p* |
| MileAge delta | | | | | | | | |
| None | | | | Reference | | | | |
| 1 | 0.020 | 0.002 | 0.037 | 0.209 | 0.017 | -0.001 | 0.034 | 0.209 |
| 2 | 0.016 | -0.007 | 0.039 | 0.352 | 0.011 | -0.012 | 0.034 | 0.485 |
| 3 | -0.011 | -0.045 | 0.023 | 0.587 | -0.019 | -0.053 | 0.016 | 0.478 |
| 4 | -0.006 | -0.056 | 0.044 | 0.804 | -0.016 | -0.066 | 0.034 | 0.587 |
| 5 | 0.084 | -0.002 | 0.170 | 0.209 | 0.067 | -0.019 | 0.154 | 0.315 |
| Metabolomic profile | | | | | | | | |
| None | | | | Reference | | | | |
| 1 | 0.085 | 0.025 | 0.144 | 0.027 | 0.053 | -0.007 | 0.112 | 0.159 |
| 2 | 0.131 | 0.048 | 0.213 | 0.020 | 0.081 | -0.002 | 0.164 | 0.139 |
| 3 | 0.057 | -0.067 | 0.181 | 0.521 | -0.017 | -0.140 | 0.107 | 0.791 |
| 4 | 0.205 | 0.023 | 0.387 | 0.091 | 0.064 | -0.118 | 0.246 | 0.566 |
| 5 | 0.299 | -0.053 | 0.650 | 0.159 | 0.117 | -0.231 | 0.466 | 0.566 |
| Frailty index | | | | | | | | |
| None | | | | Reference | | | | |
| 1 | 0.177 | 0.166 | 0.187 | <0.001 | 0.140 | 0.130 | 0.150 | <0.001 |
| 2 | 0.352 | 0.338 | 0.365 | <0.001 | 0.292 | 0.279 | 0.305 | <0.001 |
| 3 | 0.509 | 0.489 | 0.529 | <0.001 | 0.436 | 0.416 | 0.456 | <0.001 |
| 4 | 0.720 | 0.690 | 0.749 | <0.001 | 0.616 | 0.587 | 0.646 | <0.001 |
| 5 | 0.978 | 0.927 | 1.029 | <0.001 | 0.831 | 0.780 | 0.882 | <0.001 |
| Telomere length | | | | | | | | |
| None | | | | Reference | | | | |
| 1 | -0.009 | -0.020 | 0.003 | 0.242 | -0.009 | -0.021 | 0.003 | 0.242 |
| 2 | -0.001 | -0.016 | 0.015 | 0.928 | -0.002 | -0.017 | 0.014 | 0.928 |
| 3 | 0.015 | -0.007 | 0.038 | 0.268 | 0.014 | -0.009 | 0.037 | 0.278 |
| 4 | 0.033 | -0.001 | 0.067 | 0.187 | 0.029 | -0.005 | 0.063 | 0.236 |
| 5 | 0.134 | 0.075 | 0.193 | <0.001 | 0.130 | 0.072 | 0.189 | <0.001 |
| Grip strength | | | | | | | | |
| None | | | | Reference | | | | |
| 1 | 0.053 | 0.045 | 0.060 | <0.001 | 0.035 | 0.027 | 0.043 | <0.001 |
| 2 | 0.078 | 0.068 | 0.088 | <0.001 | 0.048 | 0.038 | 0.058 | <0.001 |
| 3 | 0.090 | 0.076 | 0.105 | <0.001 | 0.057 | 0.042 | 0.072 | <0.001 |
| 4 | 0.106 | 0.084 | 0.128 | <0.001 | 0.060 | 0.038 | 0.082 | <0.001 |
| 5 | 0.169 | 0.131 | 0.207 | <0.001 | 0.105 | 0.067 | 0.143 | <0.001 |

*Note:* CI = confidence interval. Model 1–adjusted for chronological age and sex; Model 2–adjusted for chronological age, sex, ethnicity, highest educational/professional qualification, annual gross household income and Townsend deprivation index. *P*-values shown are corrected for multiple testing using the Benjamini–Hochberg procedure (across exposure levels and models, separately for each ageing marker and childhood and adulthood exposures). Sample sizes reported in Table S7.

#### 10. Item-specific adverse/traumatic events in adulthood

**Table S10.** Associations between adulthood trauma items and ageing markers

| | Model 1 (adj. age and sex) | | | | Model 2 (full adjustment) | | | |
|---|---|---|---|---|---|---|---|---|
| | *β* | 95% CI | | *p* | *β* | 95% CI | | *p* |
| MileAge delta | | | | | | | | |
| Abuse, physical | -0.012 | -0.034 | 0.010 | 0.671 | -0.016 | -0.038 | 0.007 | 0.585 |
| Abuse, emotional | 0.002 | -0.015 | 0.020 | 0.996 | 0.000 | -0.018 | 0.018 | >0.999 |
| Abuse, sexual | -0.001 | -0.033 | 0.031 | >0.999 | -0.005 | -0.038 | 0.027 | 0.996 |
| Neglect, emotional | 0.013 | -0.003 | 0.029 | 0.585 | 0.008 | -0.008 | 0.024 | 0.671 |
| Economic hardship | 0.014 | -0.006 | 0.035 | 0.585 | 0.008 | -0.013 | 0.029 | 0.757 |
| Metabolomic profile | | | | | | | | |
| Abuse, physical | 0.004 | -0.077 | 0.084 | 0.925 | -0.048 | -0.128 | 0.032 | 0.425 |
| Abuse, emotional | 0.021 | -0.041 | 0.083 | 0.733 | 0.004 | -0.058 | 0.065 | 0.925 |
| Abuse, sexual | -0.008 | -0.128 | 0.112 | 0.925 | -0.069 | -0.188 | 0.050 | 0.425 |
| Neglect, emotional | 0.108 | 0.052 | 0.164 | <0.001 | 0.055 | -0.002 | 0.112 | 0.149 |
| Economic hardship | 0.194 | 0.117 | 0.271 | <0.001 | 0.117 | 0.039 | 0.195 | 0.011 |
| Frailty index | | | | | | | | |
| Abuse, physical | 0.308 | 0.295 | 0.321 | <0.001 | 0.265 | 0.252 | 0.278 | <0.001 |
| Abuse, emotional | 0.348 | 0.338 | 0.359 | <0.001 | 0.327 | 0.317 | 0.338 | <0.001 |
| Abuse, sexual | 0.387 | 0.367 | 0.406 | <0.001 | 0.344 | 0.325 | 0.363 | <0.001 |
| Neglect, emotional | 0.219 | 0.209 | 0.228 | <0.001 | 0.149 | 0.139 | 0.158 | <0.001 |
| Economic hardship | 0.290 | 0.278 | 0.303 | <0.001 | 0.209 | 0.196 | 0.222 | <0.001 |
| Telomere length | | | | | | | | |
| Abuse, physical | 0.022 | 0.007 | 0.037 | 0.019 | 0.020 | 0.005 | 0.035 | 0.025 |
| Abuse, emotional | 0.010 | -0.002 | 0.022 | 0.139 | 0.010 | -0.002 | 0.021 | 0.142 |
| Abuse, sexual | 0.019 | -0.002 | 0.041 | 0.129 | 0.022 | 0.001 | 0.044 | 0.088 |
| Neglect, emotional | -0.002 | -0.013 | 0.009 | 0.740 | -0.003 | -0.014 | 0.008 | 0.663 |
| Economic hardship | 0.022 | 0.008 | 0.037 | 0.018 | 0.017 | 0.002 | 0.031 | 0.059 |
| Grip strength | | | | | | | | |
| Abuse, physical | 0.026 | 0.016 | 0.035 | <0.001 | 0.010 | 0.000 | 0.019 | 0.048 |
| Abuse, emotional | 0.029 | 0.022 | 0.037 | <0.001 | 0.021 | 0.014 | 0.029 | <0.001 |
| Abuse, sexual | 0.035 | 0.021 | 0.049 | <0.001 | 0.017 | 0.003 | 0.031 | 0.023 |
| Neglect, emotional | 0.078 | 0.071 | 0.085 | <0.001 | 0.047 | 0.040 | 0.055 | <0.001 |
| Economic hardship | 0.085 | 0.075 | 0.094 | <0.001 | 0.051 | 0.042 | 0.060 | <0.001 |

*Note:* CI = confidence interval. Model 1–adjusted for chronological age and sex; Model 2–adjusted for chronological age, sex, ethnicity, highest educational/professional qualification, annual gross household income and Townsend deprivation index. *P*-values shown are corrected for multiple testing using the Benjamini–Hochberg procedure (across trauma items and models, separately for each ageing marker and childhood and adulthood exposures). Sample sizes reported in Tables S7.

### 11. Analytical sample sizes child and adulthood trauma

**Table S11.** Child and adulthood trauma analytical sample sizes

| Cross-classification | MileAge delta | Mortality profile | Frailty index | Telomere length | Grip strength |
|---|---|---|---|---|---|
| Neither | 21921 (32.7%) | 1446 (35.1%) | 48284 (32.6%) | 45627 (32.6%) | 48156 (32.6%) |
| Childhood | 9464 (14.1%) | 638 (15.5%) | 20745 (14.0%) | 19618 (14.0%) | 20697 (14.0%) |
| Adulthood | 17691 (26.4%) | 1053 (25.5%) | 38843 (26.3%) | 36736 (26.3%) | 38762 (26.3%) |
| Both | 17889 (26.7%) | 987 (23.9%) | 40086 (27.1%) | 37846 (27.1%) | 40018 (27.1%) |

*Note:* Numbers shown are counts and percentages.

#### 12. Adverse/traumatic events in child and/or adulthood

**Table S12.** Associations between adverse/traumatic events and ageing markers

| | Model 1 (adj. age and sex) | | | | Model 2 (full adjustment) | | | |
|---|---|---|---|---|---|---|---|---|
| | *β* | 95% CI | | *p* | *β* | 95% CI | | *p* |
| MileAge delta | | | | | | | | |
| Neither | Reference | | | | | | | |
| Childhood | 0.001 | -0.023 | 0.025 | 0.981 | 0.002 | -0.022 | 0.026 | 0.981 |
| Adulthood | 0.000 | -0.020 | 0.020 | 0.981 | -0.003 | -0.023 | 0.017 | 0.981 |
| Both | 0.032 | 0.012 | 0.051 | 0.010 | 0.028 | 0.008 | 0.048 | 0.021 |
| Metabolomic profile | | | | | | | | |
| Neither | Reference | | | | | | | |
| Childhood | 0.017 | -0.063 | 0.097 | 0.813 | 0.007 | -0.072 | 0.086 | 0.858 |
| Adulthood | 0.071 | 0.003 | 0.139 | 0.083 | 0.027 | -0.041 | 0.095 | 0.654 |
| Both | 0.135 | 0.065 | 0.204 | <0.001 | 0.083 | 0.013 | 0.153 | 0.059 |
| Frailty index | | | | | | | | |
| Neither | Reference | | | | | | | |
| Childhood | 0.213 | 0.199 | 0.227 | <0.001 | 0.206 | 0.192 | 0.220 | <0.001 |
| Adulthood | 0.199 | 0.187 | 0.211 | <0.001 | 0.154 | 0.143 | 0.166 | <0.001 |
| Both | 0.494 | 0.483 | 0.506 | <0.001 | 0.434 | 0.422 | 0.445 | <0.001 |
| Telomere length | | | | | | | | |
| Neither | Reference | | | | | | | |
| Childhood | 0.011 | -0.005 | 0.028 | 0.342 | 0.013 | -0.004 | 0.029 | 0.342 |
| Adulthood | -0.004 | -0.018 | 0.009 | 0.510 | -0.007 | -0.020 | 0.006 | 0.367 |
| Both | 0.008 | -0.005 | 0.021 | 0.342 | 0.010 | -0.004 | 0.023 | 0.342 |
| Grip strength | | | | | | | | |
| Neither | Reference | | | | | | | |
| Childhood | -0.001 | -0.011 | 0.010 | 0.899 | -0.005 | -0.016 | 0.005 | 0.399 |
| Adulthood | 0.061 | 0.053 | 0.070 | <0.001 | 0.041 | 0.032 | 0.049 | <0.001 |
| Both | 0.071 | 0.063 | 0.080 | <0.001 | 0.041 | 0.032 | 0.050 | <0.001 |

*Note:* CI = confidence interval. Model 1–adjusted for chronological age and sex; Model 2–adjusted for chronological age, sex, ethnicity, highest educational/professional qualification, annual gross household income and Townsend deprivation index. *P*-values shown are corrected for multiple testing using the Benjamini–Hochberg procedure (across exposure levels and models, separately for each ageing marker). Sample sizes reported in Table S11.

### 13. Overview of results

**Table S13.** Simplified overview of results

| | MileAge delta | Mortality profile | Frailty index | Telomere length | Grip strength |
|---|---|---|---|---|---|
| Childhood | | | | | |
| Binary | √ | ~ | √ | √ | ~ |
| Multiple[1] | √ | ~ | √ | √ | √ |
| Sum scores | √ | × | √ | √ | √ |
| Sum scores (cat.) | ~ | ~ | √ | ~ | ~ |
| Item-level | ~ | ~ | √ | ~ | ~ |
| Adulthood | | | | | |
| Binary | ~ | ~ | √ | × | √ |
| Multiple[1] | × | ~ | √ | × | √ |
| Sum scores | × | ~ | √ | √ | √ |
| Sum scores (cat.) | ~ | ~ | √ | ~ | √ |
| Item-level | × | ~ | √ | ~ | √ |
| Child and adulthood | | | | | |
| Cross-classification | ~ | ~ | √ | × | ~ |

*Note:* [1] comparison of multiple vs no exposures only. √ indicates consistent evidence of association with biological ageing marker; × indicates consistent lack of evidence of association with biological ageing marker; ~ indicates partial evidence of association with biological ageing marker (e.g., nominal association) or mixed findings.
